# Wastewater Parvovirus B19 Signal Amid Rising Maternal Cases

**DOI:** 10.1101/2025.07.07.25331044

**Authors:** Justin R. Clark, Michael J. Tisza, Alexandra L. Hammerquist, Harihara Prakash, Jessian L. Munoz, Anil Surathu, Sara J. Javornik Cregeen, Noah W. Parker, Magdalena Sanz Cortes, Blake Hanson, Jennifer Deegan, Eric Boerwinkle, Enrico R. Barrozo, Anthony W. Maresso

**Affiliations:** Department of Molecular Virology and Microbiology, Baylor College of Medicine, Houston, TX; TAILΦR Labs, Baylor College of Medicine, Houston, TX; Alkek Center for Metagenomics and Microbiome Research, Baylor College of Medicine, Houston, TX; Department of Obstetrics and Gynecology, Baylor College of Medicine and Texas Children’s Hospital, Houston, TX; Department of Obstetrics and Gynecology, Baylor College of Medicine and Texas Children’s Hospital, Division of Fetal Therapy, Houston, TX; Department of Obstetrics and Gynecology, Baylor College of Medicine and Texas Children’s Hospital, Division of Maternal-fetal Medicine, Houston, TX; The University of Texas Health Science Center at Houston (UTHealth) School of Public Health, Houston, TX

## Abstract

We report widespread detection of parvovirus B19 in Texas Wastewater using hybrid-capture virome sequencing across 43 sites. Wastewater signal correlated with clinical cases at institutional, county, and state levels and preceded case surges by one month. Full-genome coverage enabled real-time mutation tracking, highlighting wastewater’s utility for epidemiologic surveillance.

## The Study

Vertically transmitted infections are a significant threat to maternal-fetal health. Parvovirus B19 (B19V) is a small, non-enveloped DNA virus that causes Fifth Disease (erythema infectiosum), a contagious condition in children.^1^ However, infection in pregnant patients can result in hydrops fetalis, fetal anemia, and in severe cases, fetal demise. Recent surveillance reports from Europe and the United States indicate a sharp increase in B19V activity, particularly among pregnant patients and transfusion recipients. One-third of B19V-related blood transfusions in pregnant women in 2023-2024 resulted in adverse outcomes, including 22% perinatal death.^1^ B19V seroprevalence has tripled in recent years and, despite outbreaks, B19V is not tracked in national surveillance systems in the United States.^1,2^ This detection gap hinders timely public health responses and puts pregnant mothers at risk. Here, we show that B19V, like many respiratory viruses, is detectable in municipal wastewater by unbiased virome sequencing, and that wastewater signal dynamics closely parallel clinical case trends across multiple scales. In addition to correlating with outbreak timing, wastewater reads covered the full B19V genome, enabling real-time monitoring of viral mutations during the 2024 case surge.

As part of the Texas Epidemiology and Public Health Institute (TEPHI), we conduct weekly to biweekly hybrid-capture sequencing of wastewater across 43 sites and 15 cities in Texas (TexWEB: https://dashboard.tephi.texas.gov).^3–5^ Sequencing reads were aligned to the B19V J35 reference (Genbank: AY386330.1), and abundance of B19V was calculated as reads per kilobase per million filtered reads (RPKMF). We evaluated correlations between monthly RPKMF and clinical B19V case counts at three geographic scales: (i) molecularly confirmed cases at Texas Children’s Hospital (TCH), (ii) all B19V diagnoses in Harris County from the EPIC system, and (iii) all cases statewide in Texas from the EPIC system.

Spearman’s rank correlation was used to calculate correlation between wastewater and clinical datasets (Shapiro-Wilks p < 0.001 for each).

B19V was first detected at low levels in Houston-area wastewater in late 2022. The signal intensified substantially in 2024, with all 20 Houston-area monitored sites showing B19V reads by May (**Figure 1A**). In parallel, molecularly confirmed B19V cases at TCH increased 53% overall and 227% among pregnant individuals between 2023 and 2024 (p=0.00091, χ^2^ test). Overlaying monthly RPKMF and clinical data revealed a visible temporal alignment (**Figure 1B**), particularly in spring and fall 2024. The unshifted correlation was statistically significant (ρ = 0.37, p = 0.031), and became stronger when the wastewater signal was advanced by one month (ρ = 0.59, p = 0.0002; **Figure 1C**). A formal cross-correlation analysis confirmed that the peak association occurred at a -1 month lag (**Figure 1D**), consistent with wastewater leading clinical case detection. These analyses are based on a high-confidence dataset of chart-reviewed, PCR-or IgG/IgM-confirmed cases from a single institution, providing a robust foundation for broader comparisons.

**Figure 1.**
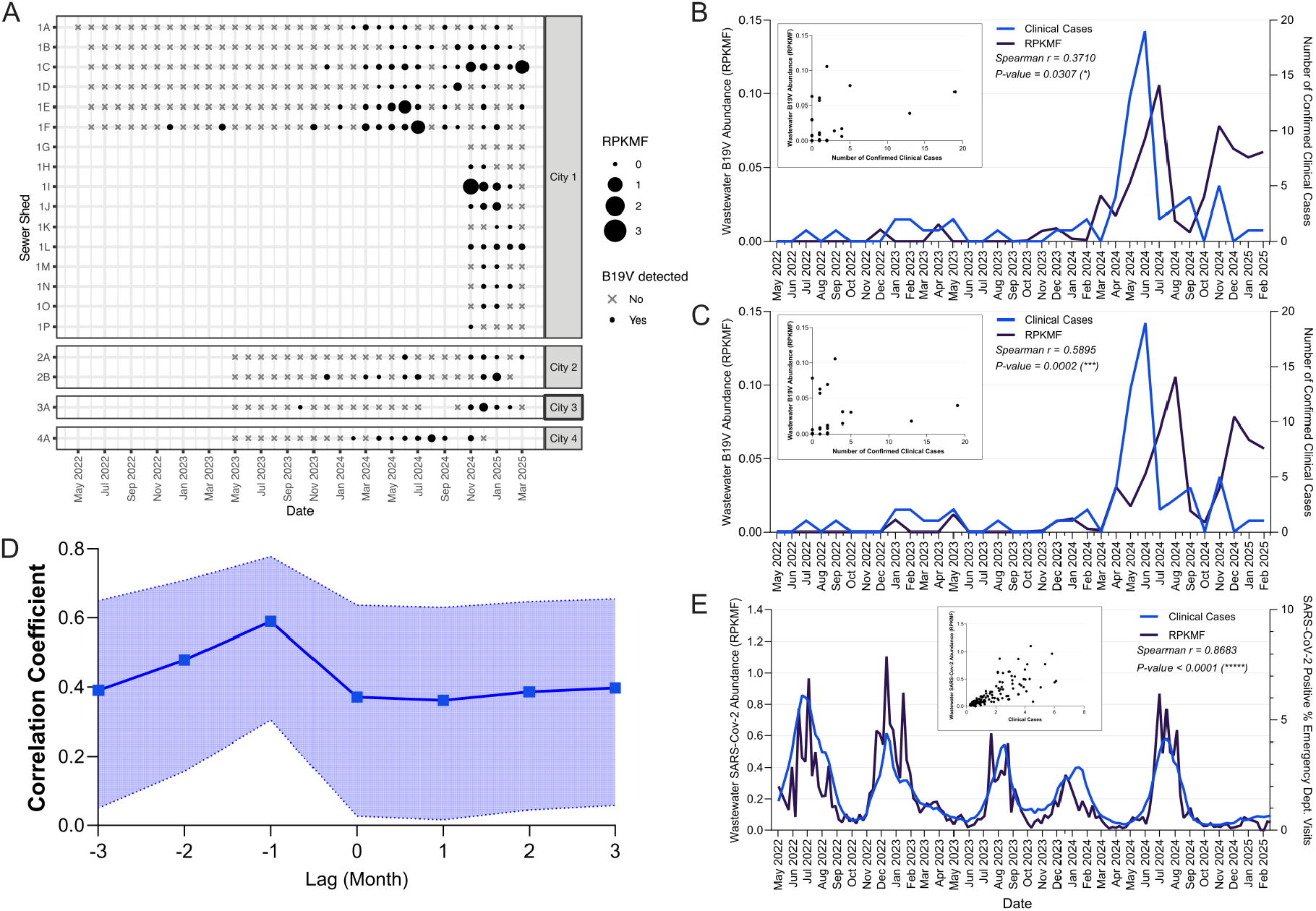
Temporal dynamics of Parvovirus B19 (B19V) in Houston-area wastewater, correlation with clinical cases, and comparison with SARS-CoV-2, and. (**A**) B19V abundance in Houston-area wastewater treatment plants from May 2022 to February 2025, measured using wastewater virome sequencing. Monitoring began at different times across sites: Site 1A in May 2022, Sites 1B–1F in July 2022, Sites 1G–1P in November 2024, and Sites 2A–4A in May 2023. Viral abundance is reported as Reads Per Kilobase per Million Filtered reads (RPKMF) (**B**) Monthly clinical cases of B19V from the Texas Children’s Hospital patient population were compared with wastewater virome sequencing data from Houston, TX area cities. **Inset**: Spearman correlation scatter plot (ρ = 0.371, P = 0.031) (**C**) Same data as in (B) with the wastewater signal advanced by 1 month (lag = -1), demonstrating improved concordance. **Inset**: Spearman correlation scatter plot (ρ = 0.574, P = 0.0004). (**D**) Cross-correlation analysis for -3 to 3 months of B19V wastewater data vs clinical cases as measured by Spearman’s rank-order correlation. Shaded bands indicate 95% confidence intervals. Shapiro-Wilk test for normality indicated data is non-normally distributed, justifying the use of Spearman rank-order statistic. The strongest correlation occurs when wastewater leads clinical cases by 1 month. (**E**) SARS-CoV-2 wastewater abundance (RPKMF) and clinical abundance (Percent positive tests for emergency department visits) in the Houston-area measured on a weekly basis, included for benchmarking. **Inset**: Spearman correlation scatter plot (ρ = 0.8683, P < 0.0001).

To benchmark our analytic approach, we previously applied the same methods to SARS-CoV-2 wastewater signal and emergency department positivity rates from the same sites and time period. That analysis yielded a strong correlation (ρ = 0.87, p < 0.0001), validating our correlation framework and sequencing pipeline (**Figure 1E**). Like many respiratory viruses, B19V is primarily transmitted via respiratory droplets, but it can also be transmitted via blood and is shed in the feces and urine of some patients— particularly during early infection. The observed lead time of ∼1 month between wastewater signal and confirmed B19V diagnoses may reflect this early shedding phase or capture subclinical infections that do not present for testing.

We next evaluated whether this temporal association extended to larger clinical datasets. Using deidentified EPIC Cosmos records encompassing over 200 healthcare facilities in Harris County, we compared B19V RPKMF values from Houston-area wastewater to B19V clinical diagnoses. The correlation was strong without any lag (ρ = 0.8192, p 0.0001; **Figure 2A**) and improved slightly with a 1-month lead time for the wastewater signal (ρ = 0.8330, p < 0.0001; **Figure 2B**). At the state level, we observed similar results.

**Figure 2.**
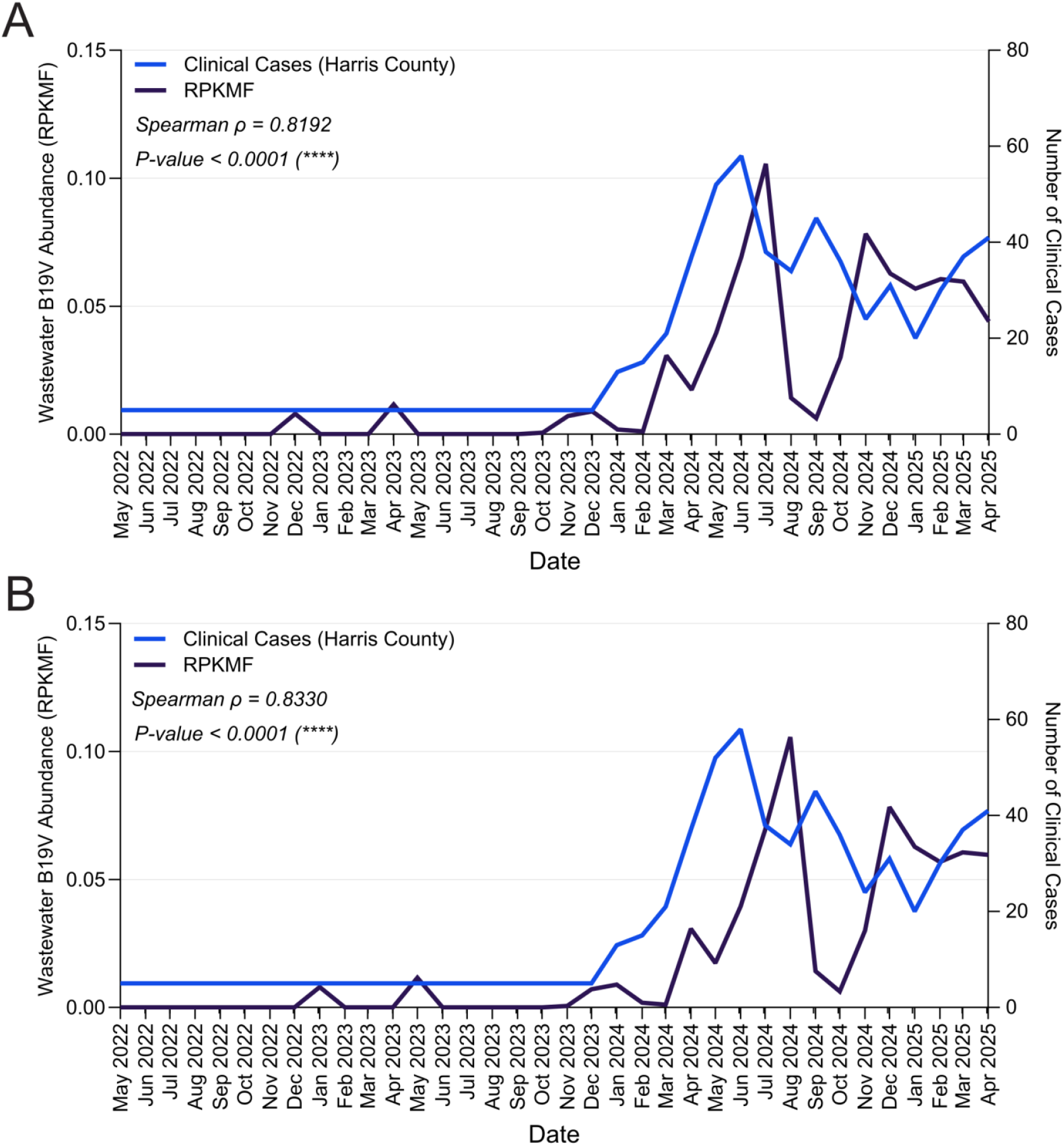
Temporal association between wastewater B19V signal and clinical cases in Harris County, Texas. (**A**) Monthly wastewater B19V relative abundance (RPKMF; purple) from Houston-area sites (Houston, Missouri City, Baytown) and monthly clinical B19V cases (blue) reported through the EPIC system for Harris County. (**B**) Same data as in (A), with wastewater signal advanced by 1 month to evaluate lead time.Spearman correlation coefficients (ρ) and P-values are shown. The peak correlation occurs when wastewater signal precedes clinical cases by 1 month, consistent with early environmental detection seen in other viruses. Note that EPIC-reported monthly case counts before January 2024 were low; when values were reported as “<10”, a value of 5 was imputed to preserve continuity.

Unshifted B19V RPKMF from Texas-wide wastewater sampling correlated with EPIC case counts across the state (ρ = 0.7780, p < 0.0001; **Figure 3A**), rising to ρ = 0.8556 (p < 0.0001) when the wastewater signal was advanced by one month (**Figure 3B**). These findings are consistent with prior studies of respiratory and enteric viruses in wastewater, where viral signal often precedes clinical diagnoses by 1-3 weeks.

**Figure 3.**
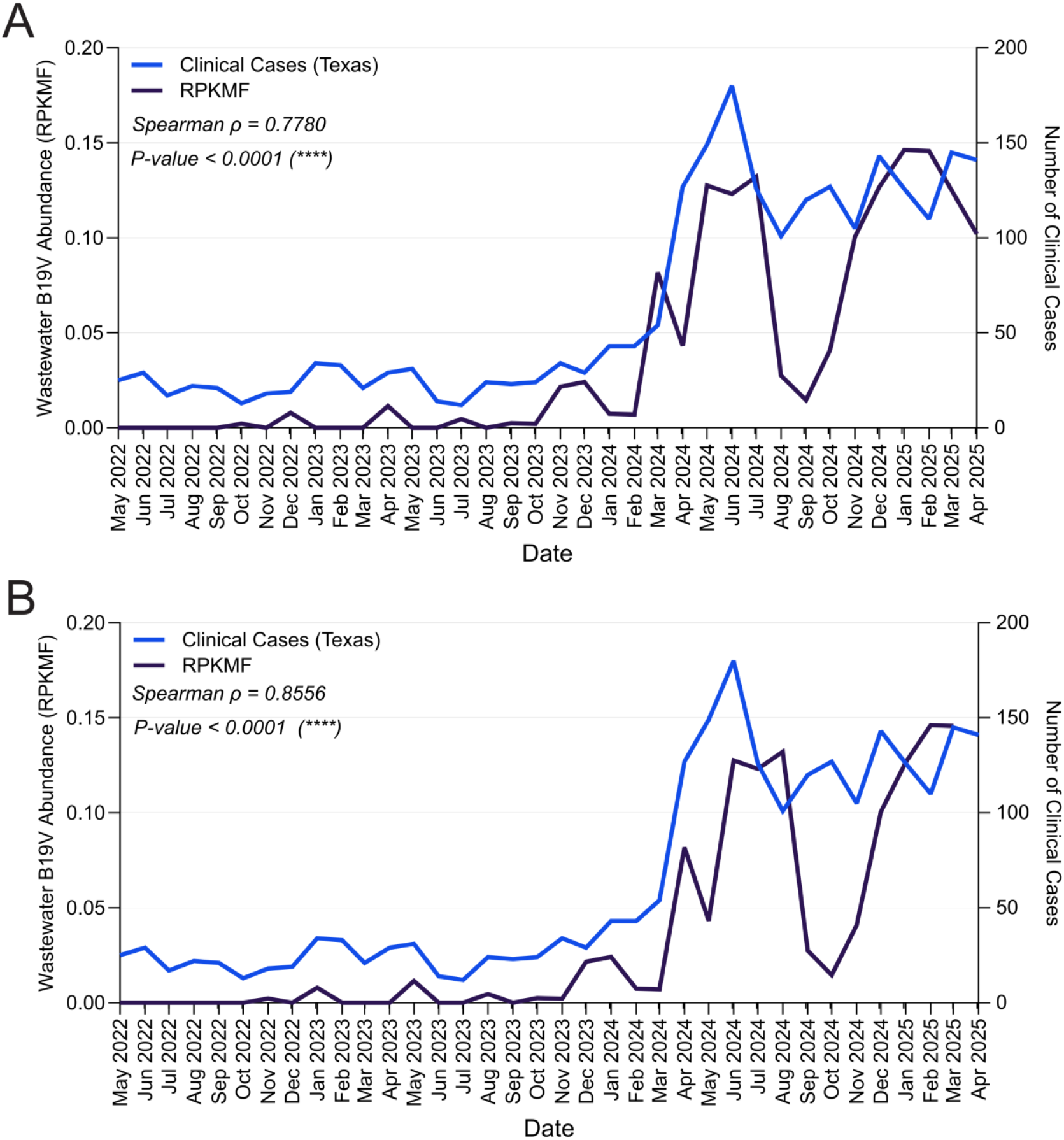
Association between statewide wastewater B19V signal and clinical case counts across Texas. (**A**) Monthly B19V abundance (RPKMF; purple) from wastewater samples collected across 15 Texas cities and statewide B19V cases (blue) reported through the EPIC clinical database. (**B**) Same data as in (A), with the wastewater signal advanced by 1 month. Correlation between wastewater and clinical data strengthens when the wastewater signal leads, suggesting a consistent early warning signal across geographic scales.

Wastewater samples also yielded high-quality B19V genome coverage, enabling mutation tracking. Using composite datasets from pre-outbreak (May 2022-Dec 2023) and post-outbreak (Jan 2024 onward) periods, we compared amino acid-altering variants recovered from hybrid-capture sequencing. Short reads spanned the full 5.6-kb reference genome with an average depth of >300x (**Figure 4A**). A total of 34 nonsynonymous mutations were observed across the NS1, VP1, VP2, and 11-kDa proteins (**Figure 4B; Appendix Table**). Several changes—most notably K10E and V88L in VP1—were undetected or rare before the 2024 surge but rose sharply during the outbreak. All reported mutations passed strand-bias and quality filtered and were supported by high read depth. These results demonstrate the feasibility of genome-resolved B19V monitoring from wastewater, including real-time variant tracking.

**Figure 4.**
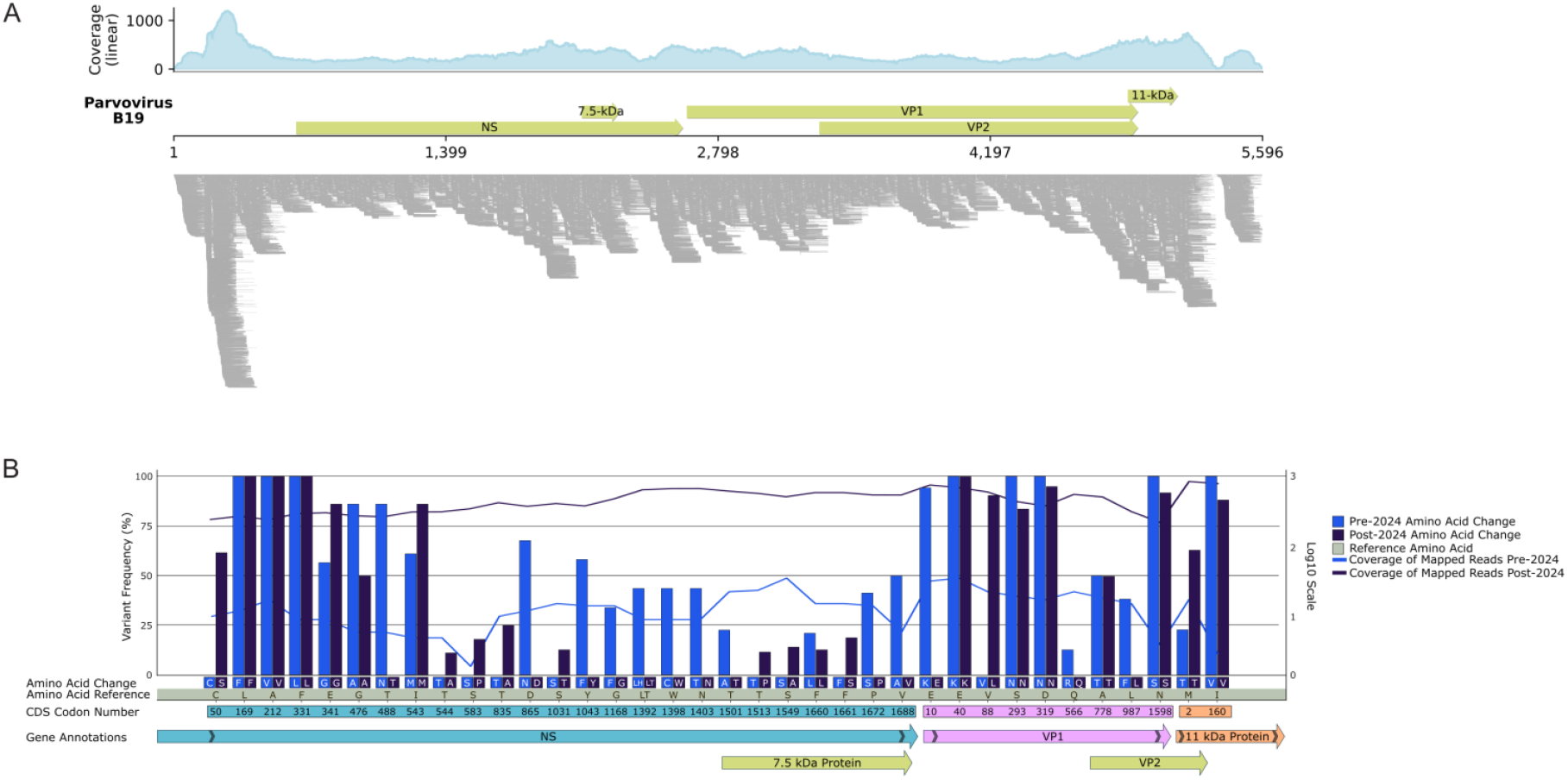
Genome-wide coverage of wastewater derived B19V reads. (**A**) Genome-wide distribution of Parvovirus B19 reads recovered from Houston-area wastewater metagenomic sequencing. The x-axis shows nucleotide position along the B19V reference genome (GenBank AY386330; total length = 5,596 nt). (**Top track**) Depth of coverage (linear scale) obtained after mapping hybrid-capture virome reads from a composite set of Houston-area wastewater influent samples (Geneious Assembler). (**Middle track**) Canonical B19V open-reading frames: the large non-structural protein (NS), 7.5-kDa accessory protein, capsid proteins VP1 and VP2, and the 11-kDa protein; coordinates are indicated beneath the scale bar. (**Bottom track**) Individual short-read alignments, illustrating even genomic coverage and the absence of large dropouts. Continuous coverage across almost the entire 5.6-kb genome confirms that near-complete B19V genomes are present in Houston-area wastewater and demonstrates the effectiveness of the hybrid-capture protocol for recovering low-abundance parvoviral sequences. (**B**) Amino acid changes detected in B19V genomes from composite wastewater samples collected before (May 2022-December 2023) and after (January 2024—April 2025) the 2024 surge. Bars indicate the frequency of nonsynonymous amino acid substitutions at each codon position (light blue = pre-2024; dark blue = post-2024). Overlaid lines show log-transformed read depth at each position in pre- and post-2024 datasets. Several variants, including K10E and V88L in the VP1 region, increase substantially during the outbreak.

## Conclusions

Several features of B19V complicate traditional surveillance. Clinical testing is typically ordered only in symptomatic pregnant patients or in response to fetal anomalies, transfusion reactions, or anemia.

Consequently, many infections likely go undiagnosed. Although B19V seroprevalence is increasing^2^, national reporting is inconsistent, and no centralized U.S. system tracks its circulation. The virus’ association with fetal morbidity and mortality raise concerns about delayed diagnosis, maternal-fetal susceptibility, and evolving pathogenesis.^1,6^ Wastewater offers an unbiased and scalable alternative. In this study, the B19V signal was consistently detected and tracked at institutional, county, and state levels, with evidence of it being a leading indicator at all three scales.

The hybrid-capture virome approach used in TexWEB also allows broad, primer-independent recovery of low-abundance viral genomes. This is particularly important for non-enveloped viruses like B19V, which may be underrepresented in standard virome pipelines or missed entirely by targeted PCR due to high sequence diversity of circulating strains. The detection of novel or shifting variants in VP1 region during the 2024 surge raises the possibility of evolving capsid epitopes or altered tropism—possibilities that warrant further investigation.

This study does have limitations. EPIC data from Harris County and the state may include coding inconsistencies or delayed reporting, although the patterns were consistent with molecularly confirmed trends from TCH. We did not evaluate seroprevalence directly, and the functional consequences of observed mutations remain unknown. Additionally, while wastewater signal appears to precede clinical reports, we were limited to more granular monthly resolution and formal predictive modeling was not performed here and should be explored in future work.

Nonetheless, these findings show that B19V is readily detectable in wastewater, that signal correlates with disease burden across geographic scales, and that sequencing enables genomic surveillance without clinical sampling. The one-month lead time observed here could be used to increase testing in maternal-fetal clinicals, prepare blood product screening, or alert providers in high-risk areas. As viral surveillance infrastructure expands, pathogens like B19V—though often overlooked—can be monitored with existing platforms. Early wastewater-based detection may help prevent fetal loss and transfusion-associated infection by prompting earlier awareness, diagnosis, and intervention.

## Data Availability

All data produced in the present study are available upon reasonable request to the authors.

## Acknowledgements

This study was supported in part by funding to TEPHI as part of the 87th legislative session (Bill SB1780), a Seed Award from Texas Children’s Hospital (ERB), and a Loan Repayment Program Award in Pediatrics (NIAID-1L40AI171990-01 to ERB). The funders had no role in study design, data collection and analysis, decision to publish, or manuscript preparation. The authors thank the Houston Health Department (Loren Hopkins, Kaavya Domakonda, and Rebecca Schneider) and Harris County Public Health for helping secure wastewater samples.

## Author contributions

Experimental design (ERB, AWM), data curation (ALJ, MJT, JRC, HP, SJJC, NWP), data analysis (ALJ, MJT, AS, JRC, SJJC, NWP), interpretation (ALJ, MJT, JRC, SJJC, JLM, NWP, MDSC, ERB, JD, AWM), writing the manuscript (ALJ, ERB, MDSC, AWM), manuscript revisions (ALJ, MJT, JRC, SJJC, JLM, NWP, AS, MDSC, AWM, ERB), and funding (ERB, AWM).

## Figure Legends

**Appendix Table 1.** Single-nucleotide variants (SNVs) in Parvovirus B19 genomes recovered from Houston-area wastewater before and after the 2024 Parvovirus Spike. Short reads from the pre-2024 composite dataset (July – Dec 2023) and the post-2024 composite dataset (Jan – Mar 2024) were mapped to the B19V reference genome (AY386330) and interrogated with the Geneious Prime Variant Finder with settings to detect variants with >10% frequency and strand bias P-value > 0.05. The table lists only sites that produce an amino-acid change in at least one period. **Column descriptions** **Position**: coordinate in the reference genome **CDS**: affected open-reading frame (NS = non-structural; VP1/VP2 = capsid; 11 K = 11 kDa protein) **Codon #**: codon position within the ORF **AA Ref / Change (Pre-2024 & Post-2024)**: reference residue and observed substitution **Freq (Pre-2024 & Post-2024)**: proportion of reads supporting the alternate allele **Cov (Pre-2024 & Post-2024)**: total read coverage depth at the position **P-Value (Pre-2024 & Post-2024)**: probability that the observed support for the variant could arise from sequencing error given the base-call qualities (variants with P-value > 0.05 were excluded)

| Position | CDS | Codon # | AA Ref | Pre-2024 | Post-2024 | Freq Pre | Freq Post | Cov Pre | Cov Post | P-value Pre | P-value Post |
| --- | --- | --- | --- | --- | --- | --- | --- | --- | --- | --- | --- |
| 665 | NS | 50 | C | - | S | - | 60.90% | 10 | 248 | - | 0 |
| 784 | NS | 169 | L | F | F | 100.00% | 100.00% | 12 | 276 | 2.50E-40 | 0 |
| 827 | NS | 212 | A | V | V | 100.00% | 99.20% | 17 | 246 | 7.90E-57 | 0 |
| 946 | NS | 331 | F | L | L | 100.00% | 100.00% | 9 | 299 | 2.50E-31 | 0 |
| 956 | NS | 341 | E | G | G | 55.60% | 83.40% | 9 | 308 | 4.00E-17 | 0 |
| 1091 | NS | 476 | G | A | A | 83.30% | 50.20% | 6 | 279 | 1.90E-18 | 0 |
| 1103 | NS | 488 | T | N | - | 83.30% | - | 6 | 270 | 1.90E-18 | - |
| 1158 | NS | 543 | I | M | M | 60.00% | 84.50% | 5 | 316 | 7.90E-11 | 0 |
| 1159 | NS | 544 | T | - | A | - | 10.10% | 5 | 317 | - | 4.80E-72 |
| 1198 | NS | 583 | S | - | P | - | 16.00% | 2 | 349 | - | 3.00E-131 |
| 1450 | NS | 835 | T | - | A | - | 25.60% | 10 | 425 | - | 2.00E-289 |
| 1480 | NS | 865 | D | N | - | 66.70% | - | 12 | 378 | 2.00E-24 | - |
| 1646 | NS | 1031 | S | - | T | - | 13.80% | 15 | 414 | - | 3.70E-135 |
| 1658 | NS | 1043 | Y | F | - | 57.10% | - | 14 | 383 | 7.50E-27 | - |
| 1783 | NS | 1168 | G | F | - | 35.70% | - | 14 | 479 | 6.30E-16 | - |
| 2007 | NS | 1392 | LT | LH | - | 44.40% | - | 9 | 642 | 2.00E-13 | - |
| 2013 | NS | 1398 | W | C | - | 44.40% | - | 9 | 668 | 2.00E-13 | - |
| 2018 | NS | 1403 | N | T | - | 44.40% | - | 9 | 669 | 2.00E-13 | - |
| 2116 | NS | 1501 | T | A | - | 22.70% | - | 22 | 616 | 8.30E-15 | - |
| 2128 | NS | 1513 | T | - | P | - | 12.00% | 23 | 574 | - | 6.70E-159 |
| 2164 | NS | 1549 | S | - | A | - | 15.40% | 34 | 514 | - | 1.30E-198 |
| 2275 | NS | 1660 | F | L | L | 20.00% | 13.80% | 15 | 588 | 3.60E-09 | 2.50E-191 |
| 2276 | NS | 1661 | F | - | S | - | 17.00% | 15 | 588 | - | 1.10E-255 |
| 2287 | NS | 1672 | P | S | - | 42.90% | - | 14 | 545 | 1.90E-19 | - |
| 2303 | NS | 1688 | V | A | - | 50.00% | - | 6 | 544 | 1.60E-10 | - |
| 2633 | VP1 | 10 | E | K | - | 90.30% | - | 31 | 750 | 7.10E-98 | - |
| 2663 | VP1 | 40 | E | K | K | 100.00% | 99.30% | 34 | 689 | 1.60E-126 | 0 |
| 2711 | VP1 | 88 | V | - | L | - | 85.00% | 22 | 599 | - | 0 |
| 2916 | VP1 | 293 | S | N | N | 100.00% | 83.90% | 19 | 441 | 1.60E-61 | 0 |
| 2942 | VP1 | 319 | D | N | N | 100.00% | 95.30% | 17 | 383 | 2.00E-53 | 0 |
| 3189 | VP1 | 566 | Q | R | - | 13.60% | - | 22 | 555 | 1.20E-08 | - |
| 3401 | VP1 | 778 | A | T | T | 50.00% | 49.20% | 18 | 510 | 2.40E-29 | 0 |
| 3401 | VP2 | 97 | A | T | T | 50.00% | 49.20% | 18 | 510 | 2.40E-29 | 0 |
| 3610 | VP1 | 987 | L | F | - | 40.00% | - | 15 | 319 | 3.20E-19 | - |
| 3610 | VP2 | 306 | L | F | - | 40.00% | - | 15 | 319 | 3.20E-19 | - |
| 4221 | VP1 | 1598 | N | S | S | 100.00% | 92.90% | 4 | 225 | 1.60E-15 | 0 |
| 4221 | VP2 | 917 | N | S | S | 100.00% | 92.90% | 4 | 225 | 1.60E-15 | 0 |
| 4891 | 11 k | 2 | M | T | T | 23.50% | 67.20% | 17 | 838 | 6.00E-11 | 0 |
| 5049 | 11 k | 160 | I | V | V | 100.00% | 89.80% | 3 | 784 | 7.90E-12 | 0 |

