## Supplemental Methods for "Wastewater Parvovirus B19 Signal Amid Rising Maternal Cases"

Wastewater Samples – We have been monitoring wastewater samples on a weekly to biweekly basis (still ongoing) since May 22, 2022 from 6-20 sites in Houston and the greater Houston area. Although we monitor other cities and sites in Texas, the Houston area is of particular focus due to its status as a central metropolitan area with much international travel. Houston also represents a carryover site from a legacy program that involved monitoring COVID-19 infections since April of 2020 in wastewater.

Extraction, library preparation, sequencing – Wastewater is first decanted upon receipt and centrifuged (3,500-4,100x g for 10 min). The supernatant is subjected to a filtration step using an ion-based cellulose filter which captures viruses in the wastewater supernatant. Filters with captured viruses are then homogenized, centrifuged and nucleic acid extracted (Qiagen QIAamp VIRAL RNA Mini Kit). RNA extracts were converted to cDNA using Protoscript II/NEBNext Ultra II kits (New England Biolabs Inc.) and nucleic acid libraries constructed with the Twist Library Preparation kit (Twist Biosciences). The libraries were pooled and incubated with > million viral probes designed against all known mammalian and plant viruses, including human (The Twist Comprehensive Viral Research Panel, Twist Biosciences). Hybridization was for 16 hours at 70 degrees Celsius followed by PCR amplification of captured sequences and sequencing using an Illumina NovaSeq 6000 SP flow cell. Raw data were converted into FASTQs and demultiplexed.

Data analysis - Read processing was performed as described.^1^ Raw fastq sequences are trimmed with BBDuk, Trimmed FASTQs are mapped to human reference genome database (GCF_000001405.39) using BBMap to remove non-viral reads and processed by EsViriru (v0.2.3; <https://github.com/cmmr/EsViritu>) to align to the Virus Pathogen Database using minimap2.^2^ CoverM (https://github.com/wwood/CoverM) thresholds are set to require at least 90% average identity across 90% of the read length and consensus sequences are extracted.^3^ After addition processing, the relative abundance of the reads in the sample are summarized with TREx (Taxonomic-based Relative-Abundance Extractor), which bins and averages RPKMF (Reads Per Kilobase of reference genome)/(Million reads passing Filtering). This metric is used to quantify read number which may relate to the relative concentration of the virus in the sample. The sequencing data may have duplicates so sometimes fastp^4^ is used to de-duplicate reads. The code is a publicly available GitHub repository: <https://github.com/TEXAS-WW/measles_ww_alignment>.

Read mapping and correlation analysis - Reads classified as parvovirus were mapped to Parvovirus B19 (J35 isolate, AY386330.1) using Geneious Prime 2025. Associations between monthly wastewater RPKMF and clinical B19V cases were evaluated with Spearman’s rank correlation (GraphPad Prism v10.4.2). SARS-CoV-2 clinical testing data were downloaded from the Texas DSHS.

Cross-correlation analysis – Reads mapping to parvovirus B19 (B19V) from Houston-area wastewater treatment plants were normalized to reads per kilobase per million filtered reads (RPKMF). For each calendar month from May 2022 through February 2025, RPKMF values were averaged (arithmetic mean) across all plants sampled that month, yielding a single relative-abundance estimate per month. Pediatric B19V case counts for the same interval were retrieved from the Texas Children’s Hospital laboratory-information system (see below). Months with no sequencing run or no reported cases were recorded as zero; no imputation or smoothing was performed.

Both the wastewater RPKMF series and the clinical case-count series failed the Shapiro–Wilk test in GraphPad Prism v10.4.2 (wastewater: W = 0.526, P < .0001; clinical: W = 0.720, P < .0001). Consequently, non-parametric Spearman rank-order correlation (ρ) was used for all cross-correlation analyses.

The wastewater series was shifted relative to the clinical series in 1-month increments from –3 to +3 months (negative lags indicate wastewater leading clinical data). For each lag k, Spearman ρ_k_, two-tailed P value, and 95 % confidence interval (CI; approximation using Fisher z transformation) were calculated in Graphpad Prism (v10.4.2). Results were plotted as ρ_k_ versus lag (Figure 1D), with shaded bands denoting 95 % CIs. The lag with the greatest absolute ρ whose CI excluded zero was interpreted as the strongest temporal association. P values are descriptive and unadjusted for the seven pre-specified lags; inference focuses on effect size (ρ) and CIs.

B19V case count data - Monthly B19V cases (01/01/2020–03/31/2025) were identified from Texas Children’s Hospital electronic records (IRB H-55722) using ICD-10 (n=4,133 non-deduplicated encounters total). Of these, 69 deduplicated cases were molecularly confirmed (PCR or serum IgG/IgM clinically validated thresholds) during the period overlapping with wastewater surveillance (01/05/2022—02/28/2025).

Data Deposition - The public repository with supporting information will be available in SRA prior to publication.
